# Breast and prostate cancer risk: the interplay of polygenic risk, high-impact monogenic variants, and family history

**DOI:** 10.1101/2021.06.04.21258277

**Authors:** Emadeldin Hassanin, Patrick May, Rana Aldisi, Isabel Spier, Andreas J. Forstner, Markus M. Nöthen, Stefan Aretz, Peter Krawitz, Dheeraj Reddy Bobbili, Carlo Maj

## Abstract

**Purpose:** Investigate to which extent polygenic risk scores (PRS), high-impact monogenic variants, and family history affect breast and prostate cancer risk by assessing cancer prevalence and cancer cumulative lifetime incidence.

**Methods:** 200,643 individuals from the UK Biobank were stratified as follows: 1. carriers or non-carriers of high impact constitutive, monogenic variants in cancer susceptibility genes, 2. high or non-high PRS (90th percentile threshold), 3. with or without a family history of cancer.

Multivariable logistic regression was used to compare the odds ratio (OR) across the different groups while Cox proportional hazards models were used to compute the cumulative incidence through life.

**Results:** Breast and prostate cancer cumulative incidence by age 70 is 7% and 5% for non-carriers with non-high PRS and reaches 37% and 32% among carriers of high-impact variants in cancer susceptibility genes with high PRS.

The additional presence of family history is associated with a further increase of the risk of developing cancer reaching an OR of 14 and 21 for breast and prostate cancer, respectively.

**Conclusion:** High PRS confers a cancer risk comparable to high-impact monogenic variants. Family history, monogenic variants, and PRS contribute additively to breast and prostate cancer risk.

## Introduction

Breast and prostate cancer represent two of the most common cancers in women and men, respectively. Within the UK Biobank (UKB) cohort, breast cancer is the most prevalent cancer diagnosis in females, and prostate cancer is the most prevalent cancer diagnosis in males (https://biobank.ctsu.ox.ac.uk/~bbdatan/CancerSummaryReport.html). Along with several other factors, predisposing genetic variants (constitutional / germline variants) play a crucial role in the risk of developing breast cancer and prostate cancer.

Both breast and prostate cancer are characterized by a high heritability estimated to be around 31% for breast cancer^1^ and 58% for prostate cancer^2^. Within breast cancer cases, approximately 5-10% are monogenic forms caused by moderate to high penetrant germline variants^3^. Similarly, in prostate cancer familial subtypes following a Mendelian inheritance have been identified ^4^. Noteworthy, in 17% of patients with prostate cancer, who were referred for genetic testing, a disease-causing germline variant could be identified^5^. Breast and prostate cancer share some susceptibility genes suggesting a potential shared genetic predisposition between the two cancer types^6^. It has also been observed that family history in first-degree relatives for prostate cancer increases women’s risk of developing breast cancer by 14%^7^. Similarly, having a first-degree relative with breast cancer increases the chance to develop prostate cancer by 18%^8^, which further underpins the hypothesis of shared genetic risk factors.

Several studies have shown the crucial role of predisposing germline variants in the etiology of breast cancer: rare high-risk variants in *BRCA1* and *BRCA2*^*9*^; rare intermediate-risk variants in *PALB2, CHEK2*, and *ATM*^*10*^; and low risk for various common variants^11^. In particular, *BRCA1/2* pathogenic variants are the most commonly linked to monogenic breast cancer, usually designated as hereditary breast and ovarian cancer^3^.

Besides the risk conferred by rare damaging variants in the strongly associated genes, different genome-wide association studies (GWAS) have identified hundreds of single nucleotide polymorphisms (SNPs) associated with breast cancer risk. Although each SNP has a negligible effect size, their cumulative impact calculated as polygenic risk score (PRS) contributes significantly to the cancer risk and improves cancer risk stratification in the general population^12^. While it is well-established that both rare and common constitutive variants are associated with breast cancer, only few studies have explored their combined effect and specifically to which extent the polygenic background acts as a risk modifier of monogenic variants of breast cancer.

For instance, the BOADICEA model is a comprehensive breast cancer prediction tool incorporating *BRCA1, BRCA2, PALB2, ATM, CHEK2* variants, along with other risk factors such as family and medical history, lifestyle, and recently also PRS^13^. In a recent study, the impact of PRS for the penetrance of the breast cancer risk variants was assessed for c.1592delT (rs180177102) in *PALB2* and c.1100delC (rs555607708) in *CHEK2* in the Finnish population^14^, and for *BRCA1/2* cancer-associated variants in a previous release of UK Biobank including a smaller cohort of 49,960 individuals with exome data^15^.

Similarly, different genes are associated with the etiology of prostate cancer. Moderate to high penetrant genes most linked to prostate cancer are *BRCA1/2, ATM, CHEK2*, and *HOXB13*^16–18^. Moreover, different studies have shown that also for prostate cancer, the cumulative risk driven by the presence of common variants as summarized by PRS models is strongly associated with the cancer risk^19^. Few studies showed the effect of PRS stratification among carriers of G84E (rs138213197) in *HOXB13*^20^, and *BRCA1/2* pathogenic variant carriers^21^. However, those studies focused only on specific variants or genes.

In the present work, we compare the prevalence and the lifetime risk of breast and prostate cancer among 200,643 individuals from the UKB. Individuals were stratified between carriers and non-carriers of rare deleterious constitutive variants in moderate to high susceptibility genes (hereafter defined as high-impact variants), high and non-high PRS, and with or without family history for cancer.

## Material and methods

### Data Source

This study was performed using genetic and phenotypic data from UKB (application number 43140). UKB is a long-term prospective population-based study, and volunteers are being recruited mainly from England, Scotland, and Wales, and involves more than 500,000 participants aged between 40-69 years at recruitment. A rich variety of phenotypic and health-related information is available on each participant, for 487,410 samples genotyping data are present and for 200,643 individuals also exomes sequencing data are available. The dataset is available for research purposes, and all participants provided documented consent^22^.

### Study participants

Breast cancer cases were defined based on self-reported code of 1002 (in data field 20001), or ICD-10 code of C50.X, or ICD-9 of 174.X in hospitalization records. For prostate cancer, cases with self-reported code of 1044 (in data field 20001), or ICD-10 code of C61, D075, or ICD-9 of 185 in hospitalization records were included. The remaining samples with no other cancer diagnosis were considered as controls. Individuals of all ancestries were included in the analysis. We excluded outliers for heterozygosity or genotype missing rates, putative sex chromosome aneuploidy, and discordant reported sex versus genotypic sex. Only individuals with both genotyping and whole-exome sequencing (WES) data were included (n=200,643). Only females with breast cancer and only males with prostate cancer were included in the analysis. We excluded one from each pair of related individuals if the genetic relationship was closer than the second degree, defined as kinship coefficient > 0.0884 as calculated by the UK Biobank (https://biobank.ctsu.ox.ac.uk/crystal/crystal/docs/ukbgene_instruct.html).

### Variant selection

Annovar^23^ was used to annotate the vcf files per chromosome from the 200,643 WES data. Variant frequencies were retrieved from the Genome Aggregation Database (gnomAD)^24^, while ClinVar^25^ annotations were considered to interpret the pathogenicity of germline variants.

The following inclusion criteria were applied to select rare high-impact variants in the UKB data:

1) only variants in protein-coding regions of *BRCA1/2, CHEK2, ATM, PALB2* genes for breast cancer and *BRCA1/2, CHEK2, ATM, HOXB13* for prostate cancer were included; 2) allele frequency (AF) <0.005 in any ethnic subpopulation of gnomAD; 3) not annotated as “synonymous”, “non-frame shift deletion” and “non-frameshift insertion”; 4) annotated as “pathogenic” or “likely pathogenic” based on ClinVar. The same variant filtering approach has been applied in a recent analysis aimed at identifying monogenic variants^15^. Individuals having any of the identified variants in the moderate to high penetrant genes were classified as carriers of cancer-associated high-impact variants.

### Polygenic risk score

To generate the PRS, we used a previously validated PRS for breast and prostate cancer containing 3820 and 103 variants, respectively^21,26^. The PRS was calculated through UKB genotype data using the PLINK 2.0^27^ scoring function. We applied a previous approach to minimize variance in PRS distributions across genetic ancestries^28^. We fit a linear regression model using the first four ancestry principal components (PCs) to predict the PRS over the entire dataset (PC_PRS∼PC1 + PC2 + PC3 + PC4). The PC adjusted PRS was calculated by subtracting PC_PRS from the raw PRS and used for the subsequent association tests.

### Statistical analysis

Individuals were stratified based on the PRS percentile, presence or absence of high-impact variants, and family history. We considered family history of cancer in parents and siblings as reported by participants (Data-fields: 20110, 20107, 20111). We assigned individuals to non-high (<90% PRS) and high (>90% PRS) where the definition of a high PRS (above the 90th percentile) followed a previous study^18^. The rational to assign PRS into two risk classes (i.e., high risk vs remaining of the population) is in line with the hypothesis that PRS is associated with a non-linear increase for extremely high PRS as observed for other phenotypes^12^. Noteworthy, we observed a similar trend in our study when comparing the prevalence of breast and prostate cancer by PRS percentile (Supplementary Figure S1).

Non-high PRS, non-carrier, and an absent family history correspond to the large majority of individuals (78.8% and 81.1%, for breast and prostate cancer, respectively); therefore, this group was used as a reference to assess the cancer prevalence in the population (i.e., to compute the ORs). We performed both a gene-unspecific (i.e., carriers of variants in all susceptibility genes) and a gene-specific analysis. In addition, for breast cancer, we also stratified between high-impact variants carriers in genes characterized by moderate/intermediate penetrance (*ATM-CHEK2-PALB2*) and carriers of high-impact variants in high-risk genes (*BRCA1/2*) to assess the impact of PRS in the two groups. For each carrier group, we computed the OR using a logistic regression model adjusted for age at recruitment and the first four PCs.

We estimated the lifetime risk by age 70 resulting from monogenic variant carrier status and the PRS. We fit a Cox proportional hazards model using the R package *survival*. We used age as the time scale, representing the time-to-event, in cases (age at first assessment), and in controls (age at last visit). The adjusted model included carrier status, age, and the first four ancestry PCs, while adjusted survival curves were plotted with the R package *survminer*. For all statistical analyses, we used R 3.6.3.

## Results

### Stratification of UKB cohort individuals for cancer prevalence, family history, and genetic risk factors

Within the 200,643 UKB individuals with available genotyping and exome data, we identified 3,739 breast cancer prevalent cases with a mean age at diagnosis of 52.2 years. The remaining 85,974 women with no other cancer diagnosis were considered as controls, the mean age at last visit was 56.8 years (Table S1).

For prostate cancer, a total 1,332 prevalent cases were identified with a mean age at diagnosis of 60.3 years. The remaining 73,111 men with no other cancer diagnosis were considered as controls, the mean age at last visit was 57.0 years (Table S2).

Noteworthy, both in breast and prostate cancer, there is a significantly higher proportion of individuals with family history for cancers not only among carriers of high-impact variants (OR = 1.79 and 1.47, P < 0.01), but also among individuals with high-PRS (OR = 1.49 and 1.36, P < 0.01) (Table 1 and Table 2).

**Table 1:** Characteristics of the participants by carrier status and polygenic risk score strata in breast cancer

|  | *Noncarrier &*<br>*NonhighPRS* | *Carrier &*<br>*HighPRS* | *Carrier &*<br>*NonhighPRS* | *Noncarrier &*<br>*HighPRS* |
| --- | --- | --- | --- | --- |
| *Participants, N* | 78962 | 223 | 1780 | 8748 |
| *Cases, N (%)* | 2808 (3.56) | 48 (21.52) | 169 (9.49) | 714 (8.16) |
| *Controls, N* | 76154 (96.44) | 175 (78.48) | 1611 (90.51) | 8034 (91.84) |
| *Age\*, mean (SD)* | 56.7 (8.36) | 55.58 (8.55) | 55.56 (8.72) | 56.32 (8.35) |
| *Family history of breast cancer, n (%)* | 8301 (10.51) | 45 (20.18) | 317 (17.81) | 1309 (14.96) |

*\*Age: age at onset for cases, and age at last visit for controls.*

**Table 2:** Characteristics of the participants by carrier status and polygenic risk score strata in prostate cancer.

|  | *Noncarrier &*<br>*NonhighPRS* | *Carrier &*<br>*HighPRS* | *Carrier &*<br>*NonhighPRS* | *Noncarrier &*<br>*HighPRS* |
| --- | --- | --- | --- | --- |
| *Participants, N* | 65397 | 208 | 1602 | 7236 |
| *Cases, N (%)* | 1001 (1.53) | 21 (10.1) | 45 (2.81) | 265 (3.66) |
| *Controls, N* | 64396 (98.47) | 187 (89.9) | 1557 (97.19) | 6971 (96.34) |
| *Age\*, mean (SD)* | 57.14 (8.64) | 56.01 (8.79) | 56.2 (8.58) | 56.75 (8.59) |
| *Family history of breast cancer, n (%)* | 5028 (7.69) | 32 (15.38) | 171 (10.67) | 731 (10.1) |

\*Age: age at onset for cases, and age at last visit for controls.

### Distribution of carriers of high-impact variants within the UKB cohort

We identified 2,003 carriers of 318 pathogenic or likely pathogenic variants in the five analyzed breast cancer susceptibility genes i.e., *BRCA1/2, PALB2, CHEK2*, and *ATM*.

In addition, 1,810 carriers of 278 pathogenic or likely pathogenic variants were found in the five considered prostate cancer susceptibility genes, i.e., *BRCA1/2, ATM, CHEK2*, and *HOXB13*. Annotations of the considered variants and number of carriers are available in the Supplementary Materials (SM).

### PRS distribution within the UKB cohort

Breast and prostate cancer PRS follow a normal distribution (raw and PC-adjusted PRS are shown in Supplementary Figure S2) and are significantly higher in cases compared to controls (P < 0.01) (Supplementary Figure S3).

We observed a non-linear increase of cancer risk for individuals in the extreme right tail of the PRS distribution (Supplementary Figure S1 - disease prevalence by PRS percentile for both breast and prostate cancer). This corroborates the hypothesis that PRS can be used to stratify individuals into risk classes according to a liability threshold model^29^ (i.e., high risk and non-high risk).

### Interplay between high-impact variants and PRS

None of the high-impact variants used to define carrier status is included in PRS, thus they represent an independent genetic signal. We observed no different PRS distributions according to the carrier status (Supplementary Figure S4). As expected, carriers as well as high-PRS group (90% percentile threshold) show significantly higher cancer ORs. The highest OR is observed when both types of genetic contribution are present (Figure 1).

**Figure 1:**
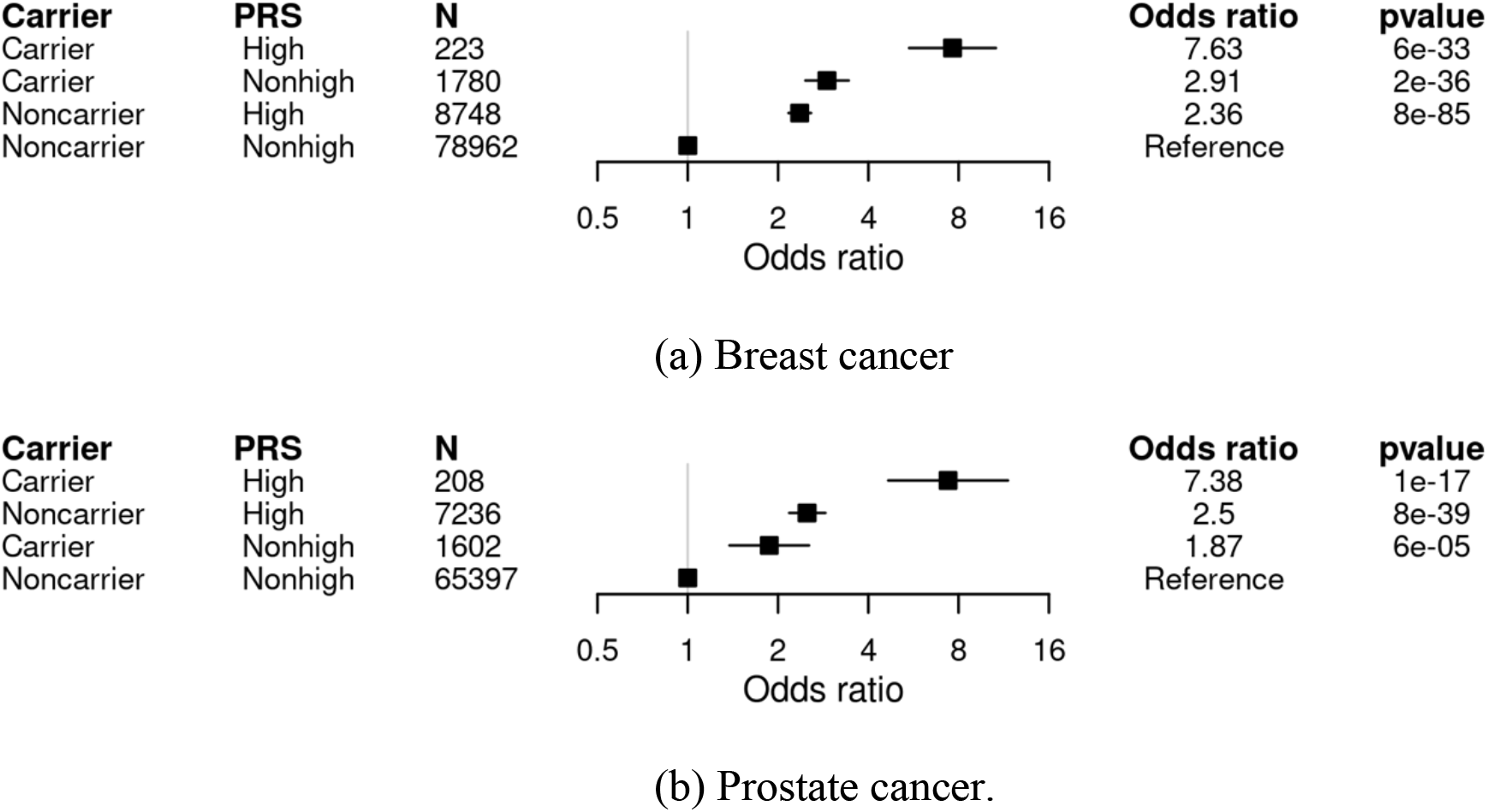
Cancer OR among individuals stratified for presence of high-impact variants and PRS. Carriers and noncarriers were categorized into two strata based on their polygenic score: Non-high (<90 % percentile), or high (>90 % percentile) PRS. The OR was calculated from a logistic regression model with age, and the first four principal components of ancestry as covariates. The reference group was noncarriers with Non-high polygenic score. The adjusted odds ratio is indicated by the black boxes. The 95% confidence intervals are indicated by the horizontal lines around the black boxes.

The cumulative incidence by age 70 is the lowest among non-carriers with non-high PRS and the highest among carriers with high-PRS, values range from 7% and 5% to 37% and 32% for breast and prostate cancer, respectively (Figure 2).

**Figure 2:**
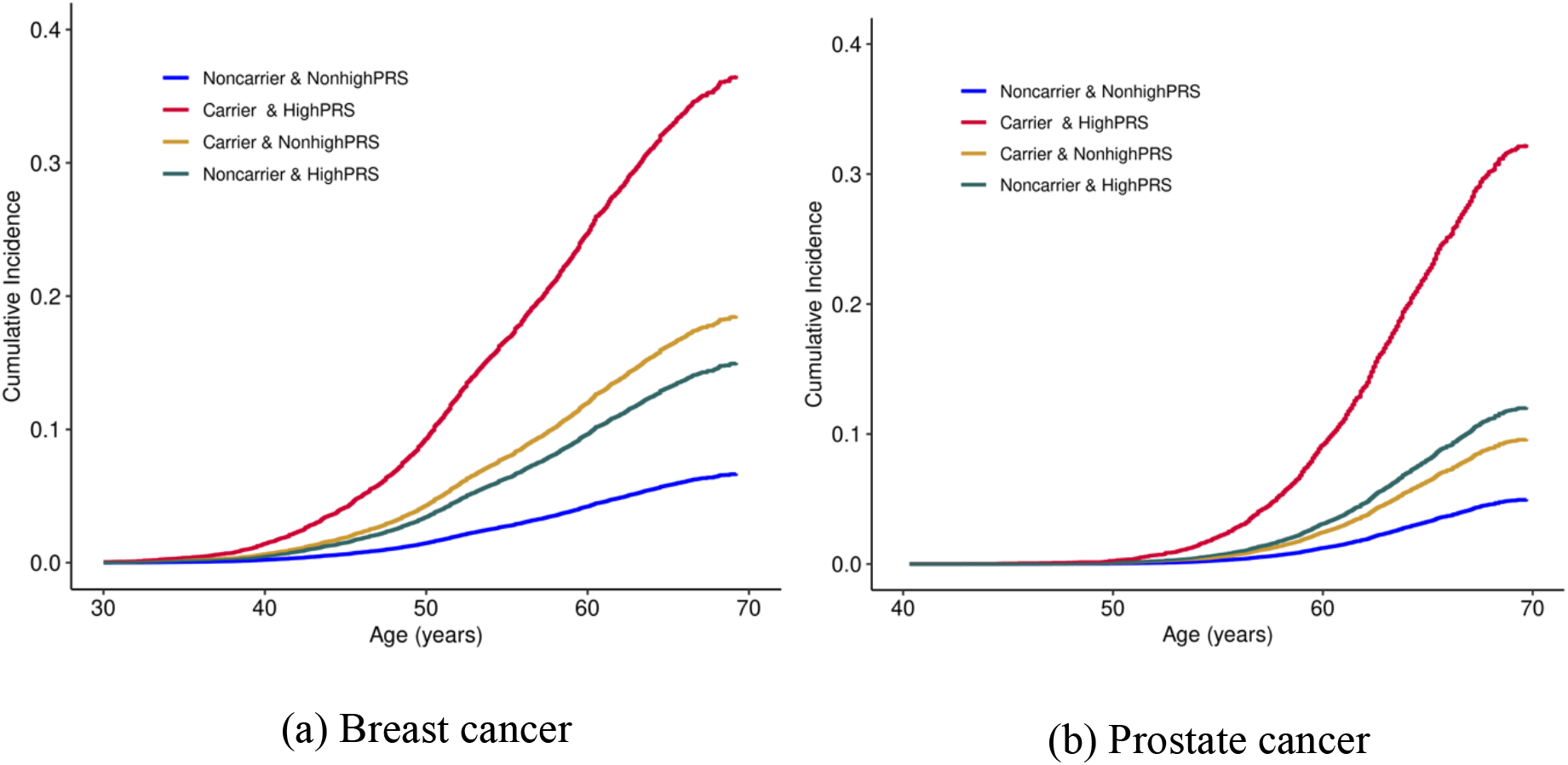
Life-time incidence of cancer among individuals stratified for the presence of high-impact variants and PRS. Cumulative incidence was estimated from a cox-proportional hazard model using age, and the first four ancestry principal components.

By testing different percentile thresholds to stratify for high and non-high PRS (from 75th to 90^th^ percentile), we observed that the higher the PRS the higher the corresponding OR (Supplementary figure S5). We did not consider higher PRS thresholds as this would have led to too few individuals to perform an analysis based on additional further stratification for family history and carriers of high-impact variants.

### The impact of polygenic risk score in single gene variant carriers

We estimated how PRS influences breast cancer prevalence among female carriers of pathogenic or likely pathogenic variants in the analyzed susceptibility genes (Supplementary Figure S6). Carriers of high-impact variants in *BRCA2* with high-PRS reached the highest OR of 9.1. For all genes except *CHEK2* being a carrier of a high-impact variant only, is associated with a higher OR compared to having only a high-PRS. However, the combined effect of being a carrier of a variant in any of the genes and having a high-PRS leads to a further increase in the OR.

Similar, having a high PRS is associated with an increased OR for prostate cancer among carriers of high-impact variants in any of the candidate risk genes (Supplementary Figure S7). The highest OR of 10.6 is reached among *HOXB13* variant carriers combined with a high PRS. However, in contrast to breast cancer, in prostate cancer ORs are higher in non-carriers with high-PRS compared to carriers of high-impact variants with non-high PRS except for the *HOXB13* gene.

### The impact of PRS in carriers of moderate/intermediate and high-risk genes variant

For breast cancer, we also performed a separate analysis for the high-risk genes *BRCA1/2* and the moderate/intermediate-risk genes *PALB2, CHEK2*, and *ATM*. We estimated how PRS influences breast cancer prevalence among female carriers of high-impact variants within the two groups. Carriers of high-impact variants in *BRCA1/2* have a higher OR than individuals with only high PRS (3.7 versus 2.4, Supplementary Figure S8a). Instead, carriers of high-impact variants in the moderate/intermediate-risk genes have a comparable OR with respect to individuals with high PRS alone (2.41 versus 2.35, Supplementary Figure S8b). However, high PRS modifies the penetrance of high-impact variants in both moderate/intermediate and high-risk genes (OR = 7.4 and 8.1, Supplementary Figure S8).

### Inclusion of family-history on the cancer risk stratification

Family history is a strong risk factor for developing cancer. Family history of cancer is present in 20.4% and 20.5% of cases and 10.7% and 7.8% of controls (OR = 2.1 and 3.1, P < 0.01) for breast and prostate cancer, respectively (Supplementary Table S1 and S2). Taking individuals with no family history and non-high PRS as a reference, both family history and PRS are associated with a higher risk (Supplementary Figure S9). The presence of both positive family history and a high PRS leads to a further increase of the cancer risk as assessed by the OR that reaches a value of 4.6 in breast cancer and 6.7 in prostate cancer (Supplementary Figure S10).

The full model including PRS, carrier status, and family history shows an additive contribution to both breast and prostate cancer risks as measured by the ORs across individuals stratified for each of the three risk factors (Figure 3).

**Figure 3:**
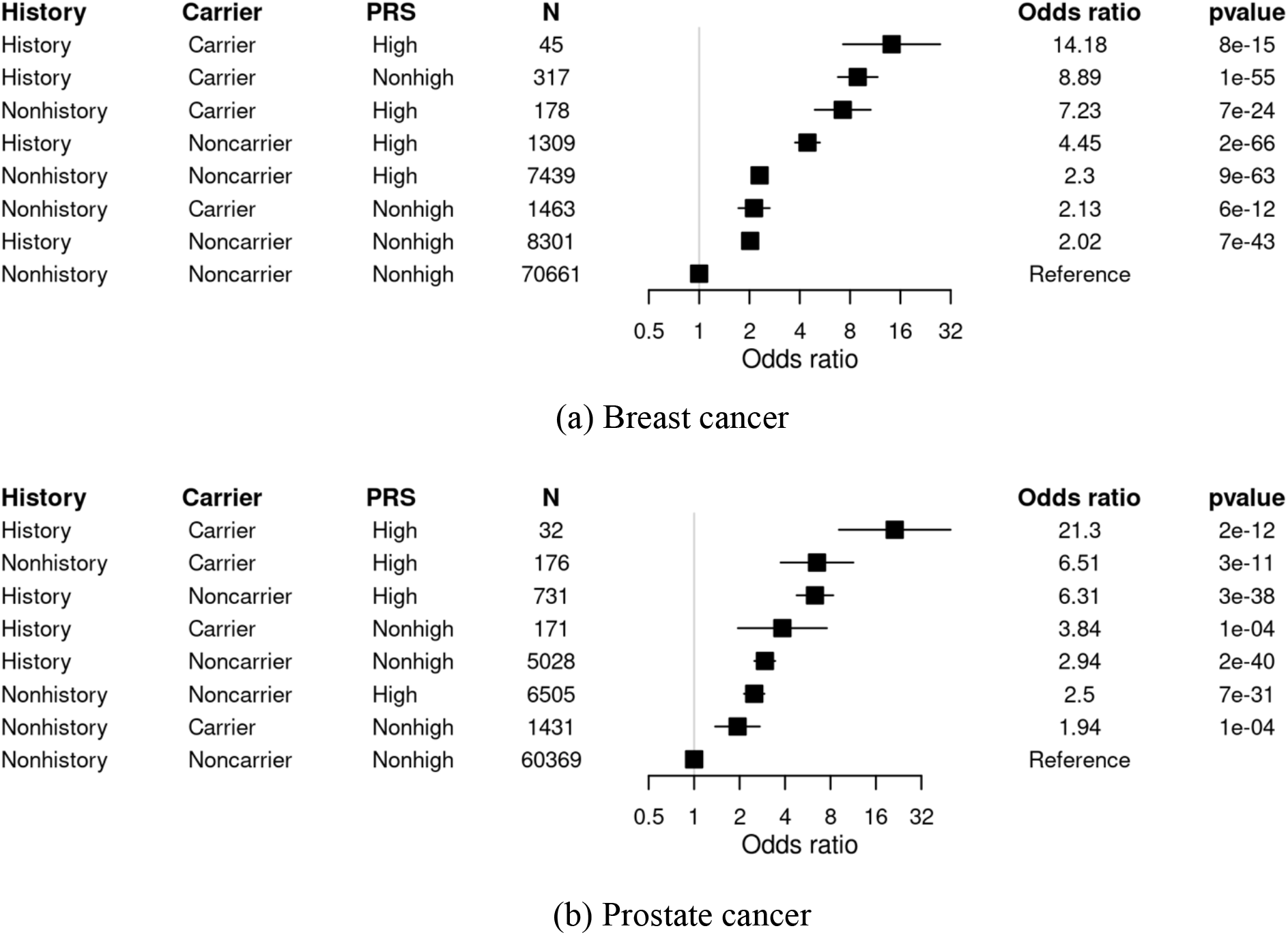
Interplay of high-impact variants, family history, and PRS. ORs for cancer was estimated from logistic models adjusted for age and first four ancestry principal components. The 95% confidence intervals are indicated by the horizontal lines around the black boxes.

## Discussion

In this study, we analyzed how breast and prostate cancer prevalence within the UKB cohort is affected by genetic susceptibility and family history. We considered both the genetic component driven by rare moderate to high penetrant variants associated with hereditary forms of cancer and the polygenic background present in all individuals.

Our results support the hypothesis of cumulative genetic risks due to both rare high-impact variants and the polygenic background. We observed a higher prevalence of cancer in the suspected hereditary forms of breast and prostate cancer in individuals with high-PRS. This result corroborates the role of polygenic background as a modifier of the breast and prostate cancer risk among high-penetrance variant carriers unselected for specific clinical criteria (UKB cohort) as observed in other studies focused on specific genes or variants^14,15^. The analysis of the corresponding age-dependent cumulative incidence of breast and prostate cancer indicates that the age of onset is also jointly influenced by the presence of high-impact variants and the polygenic contribution.

We observed that breast cancer risk conferred by only high-PRS is comparable to the risk conferred by being a carrier of a high-impact variant alone (OR respectively of 2.3 and 2.1). The same analysis for prostate cancer revealed that the risk conferred by a high PRS only is even higher than the risk conferred by being a carrier of a high-impact variant alone (2.5 and 1.9, respectively). These results suggest that a PRS can stratify the large group of non-carrier individuals similarly for breast and prostate cancer risks compared to well-known monogenic forms. According to these findings, there should be a potential benefit for both the general population and unselected at-risk individuals carrying high-penetrant variants from the inclusion of PRS in healthcare prevention policies, as risk-stratified screening might improve early disease detection and prevention^30,31^.

Moreover, with increasing population-based cohort size, the PRS can better define a small group of very high-risk non-carrier individuals in the extreme tail of the polygenic risk distribution. PRS seems to comprise cancer risks that are comparable to carriers of monogenic conditions.

In addition, our results show that the inclusion of the family history can further and independently improve the risk stratification besides the included genetic factors. Previous studies have discussed that family history is mainly associated with monogenic variants and minorly with PRS^32,33^. Our data suggest that there are additional risk factors besides of those captured by frequent low-penetrant SNPs (PRS) and rare high-impact variants. These might be common and rare structural variants (non-coding variants, large copy number variations); intermediate and low-impact variants which are not included routinely or in PRS models, and non-genetic contributors such as environmental / lifestyle factors. Our analysis shows that PRS, carriers, and family act independently and additively on breast and prostate cancer.

Our study has different limitations. First, there is evidence of a “healthy volunteers” selection bias of the UKB population, and thus the results might not be generalizable in terms of effect sizes^34^. Second, our risk assessment was based solely on genetic variants and family history and did not include other risk factors. Previous studies on UKB showed that lifestyle modifiable risk factors play a pivotal role in cancer prevalence^35^, and a shared lifestyle within families could influence family history with the disease^36^. That might explain the independent association of the family history concerning the genetic risk. Finally, although we performed the analysis on the whole UKB cohort, we could not test the risk stratification generalizability across different populations. PRS could be biased towards the European population as PRS was constructed based on European reference GWAS. Thus, PRS might be a worse predictor in non-European or admixed individuals, as previously discussed in different studies^37^.

In conclusion, we show the significant role of PRS in both the general population and carriers of monogenic, high-risk variants. A high-PRS strongly alters the penetrance of monogenic variants and influences the disease onset. The data suggest that stratification of individuals based solely on the PRS can reach ORs in line with the ones associated with rare high-penetrant variants which are currently subject to risk adapted tailored surveillance programs. As a consequence, PRS can define a high-risk group within the general population which could be considered as subjects for intense surveillance measures similar to those offered to carriers of monogenic forms. These findings highlight the potential usefulness of PRS in the context of cancer risk stratification. Our analysis also suggests that family history for breast and prostate cancer represents an additional independent risk factor to the genetic data which contributes additively to the disease risk.

## Data Availability

Genome-wide genotyping data, exome-sequencing data, and phenotypic data from the UK Biobank are available upon successful project application.

https://www.ukbiobank.ac.uk/

## Acknowledgements

UK Biobank analyses were conducted via application 52446 using a protocol approved by the Partners HealthCare Institutional Review Board. P.M was supported by the Luxembourg National Research Fund (FNR) as part of the National Centre of Excellence in Research on Parkinson’s disease (NCER-PD, FNR11264123) and the DFG Research Units FOR2715 (INTER/DFG/17/11583046) and FOR2488 (INTER/DFG/19/14429377). CM and EH are supported by the BONFOR-program of the Medical Faculty, University of Bonn (O-147.0002). SA and IS are members of the European Reference Network on Genetic Tumor Risk Syndromes (ERN GENTURIS) -Project ID No 739547. The authors acknowledge the use of de.NBI cloud and the support by the High Performance and Cloud Computing Group at the Zentrum für Datenverarbeitung of the University of Tübingen and the Federal Ministry of Education and Research (BMBF) through grant no 031A535A.

## Contributions

Conceptualization: EH, DRB and CM. Analysis: EH; supervision of the work: PK, PM, DRB and CM; results interpretation: AF, IS, SA, MMN and PK; Writing, review and editing: EH, PM, RA, IS, SA, MMN, PK, DB and CM.

## Competing interests

The authors declare no conflicts of interest

## Notes

### Competing Interest Statement

The authors have declared no competing interest.

### Funding Statement

Patrick May was supported by the Luxembourg National Research Fund (FNR) as part of the National Centre of Excellence in Research on Parkinson disease (NCER-PD/FNR11264123) and the DFG Research Units FOR2715 (INTER/DFG/17/11583046) and FOR2488 (INTER/DFG/19/14429377). Carlo Maj and Emadeldin Hassanin are supported by the BONFOR-program of the Medical Faculty University of Bonn (O-147.0002). Stefan Aretz and Isabel Spier are members of the European Reference Network on Genetic Tumor Risk Syndromes (ERN GENTURIS) -Project ID No 739547. The authors acknowledge the use of de.NBI cloud and the support by the High Performance and Cloud Computing Group at the Zentrum for Datenverarbeitung of the University of Tübingen and the Federal Ministry of Education and Research (BMBF) through grant no 031A535A.

### Author Declarations

all ethical guidelines have been followed

