## Supplementary for "Breast and prostate cancer risk: the interplay of polygenic risk, high-impact monogenic variants, and family history"

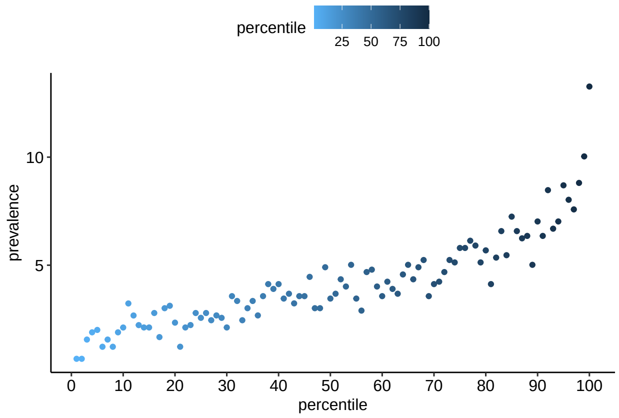


(a) Breast cancer

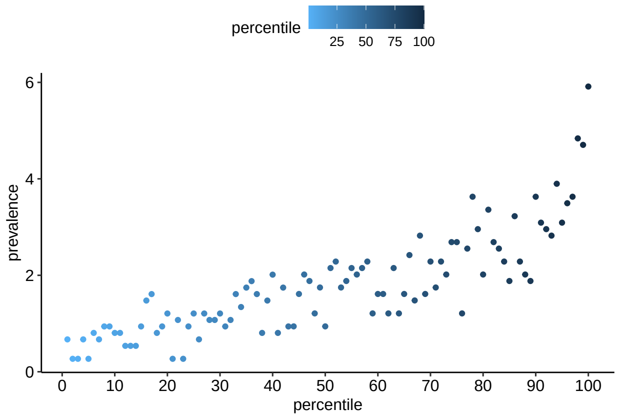


(b) Prostate cancer

Figure S1: Prevalence of the breast and prostate cancer according to polygenic risk score (PRS) perecentiles.

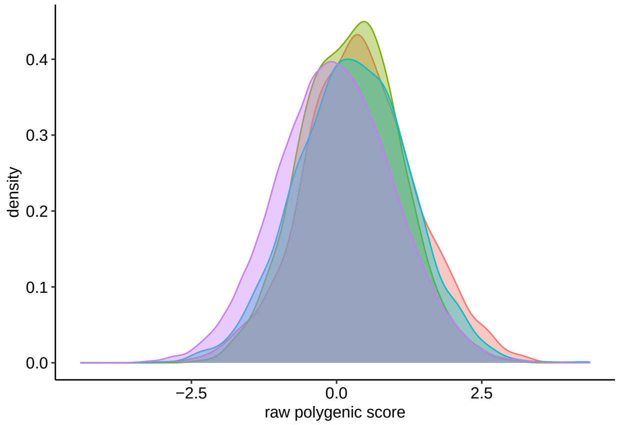


(a) Breast cancer

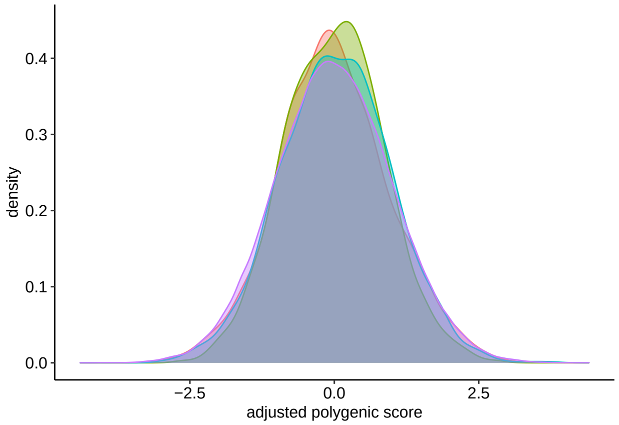


(b) Breast cancer

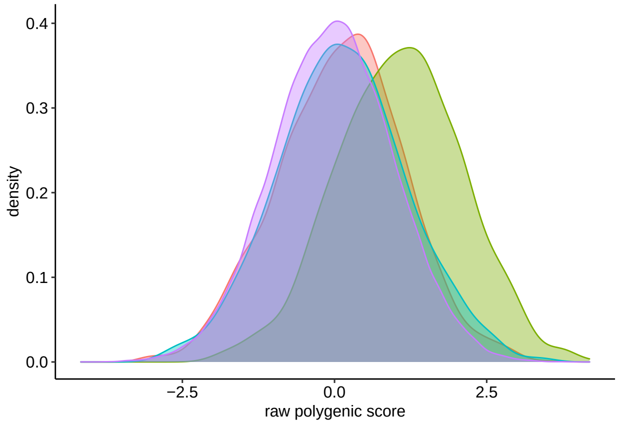


(c) Prostate cancer

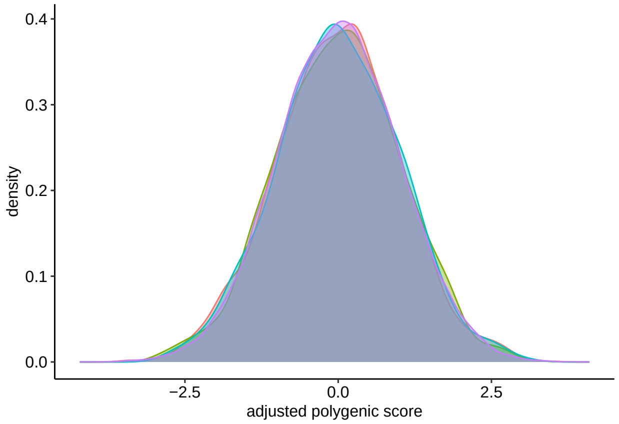


(d) Prostate cancer

Figure S2: Distributions of the breast and prostate cancer PRS across self-reported ethnic groups in the UK Biobank cohort (Data-Field 21000). Raw PRS was adjusted using first four principal components. Distributions of: a) raw PRS in breast cancer; b) adjusted PRS in breast cancer; c) raw PRS in prostate cancer; d) adjusted PRS in prostate cancer.

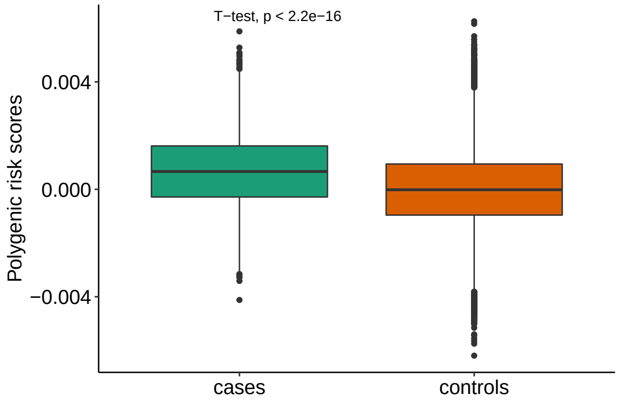


(a) Breast cancer

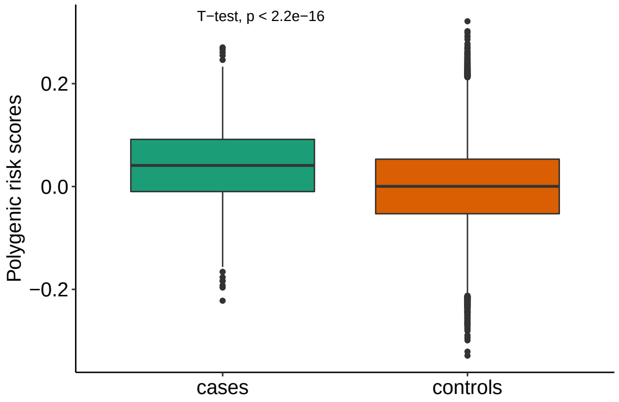


(b) Prostate cancer

Figure S3: PRS among breast and prostate cases versus controls.

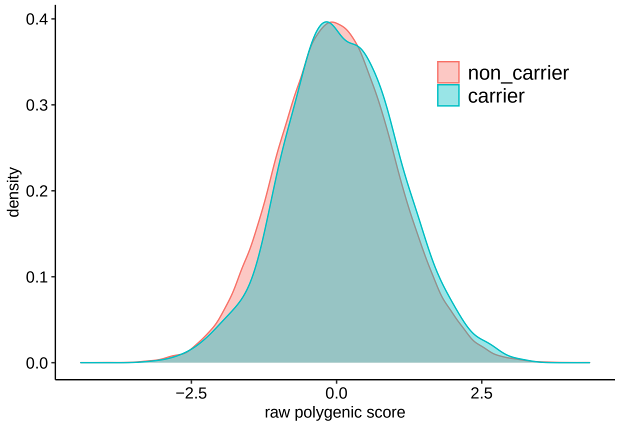


(a) Breast cancer

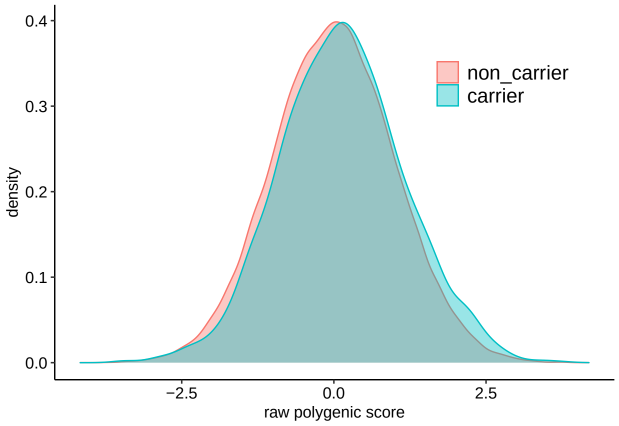


(b) Prostate cancer

Figure S4: Distributions of the breast and prostate cancer PRS across carrier status in the UK Biobank cohort.

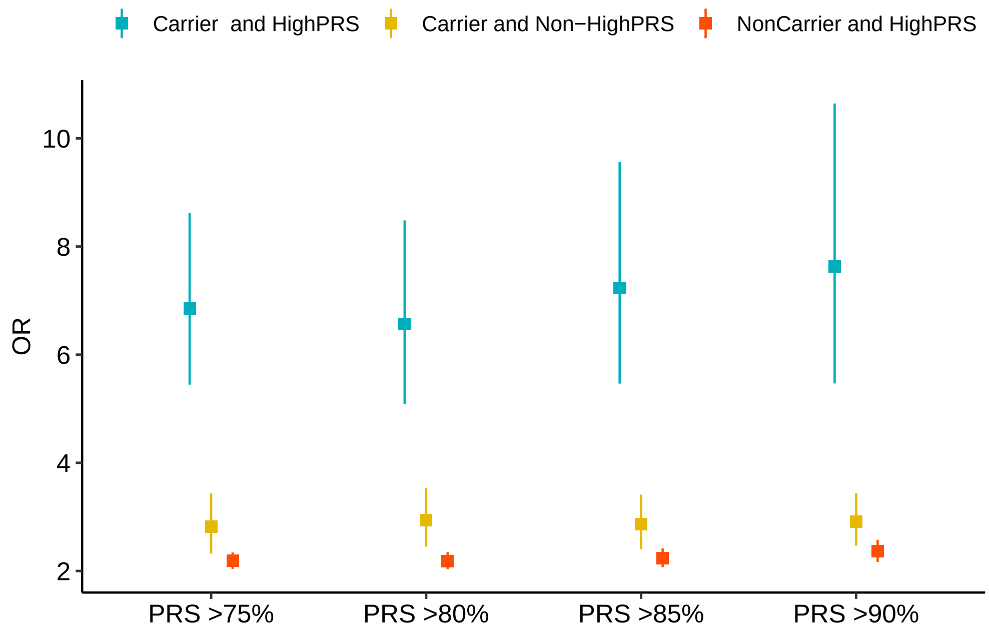


(a) breast cancer

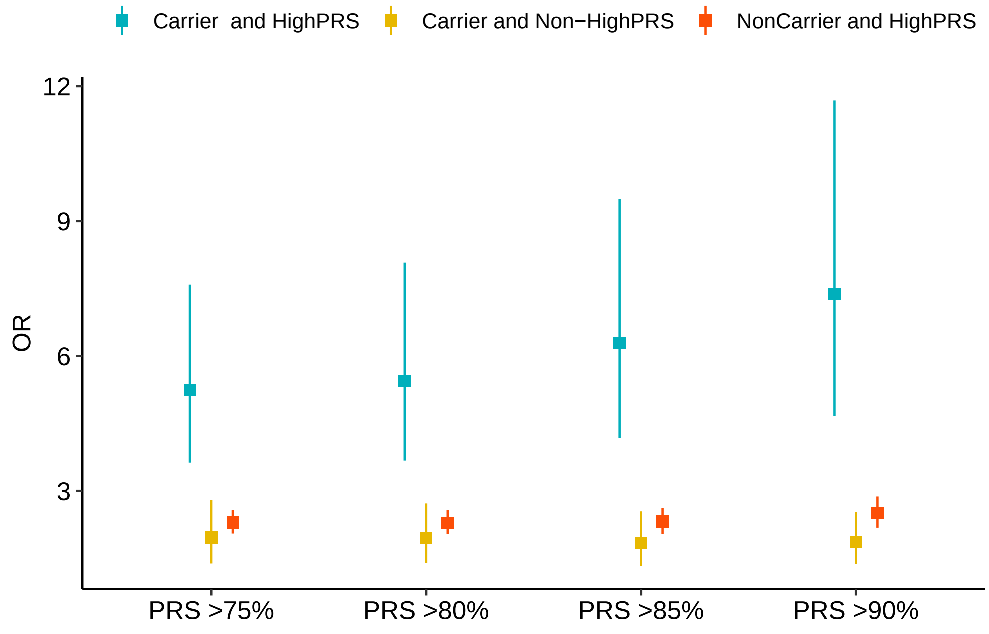


(b) prostate cancer

Figure S5: Impact of PRS across individulas with monogenic variants carriers.

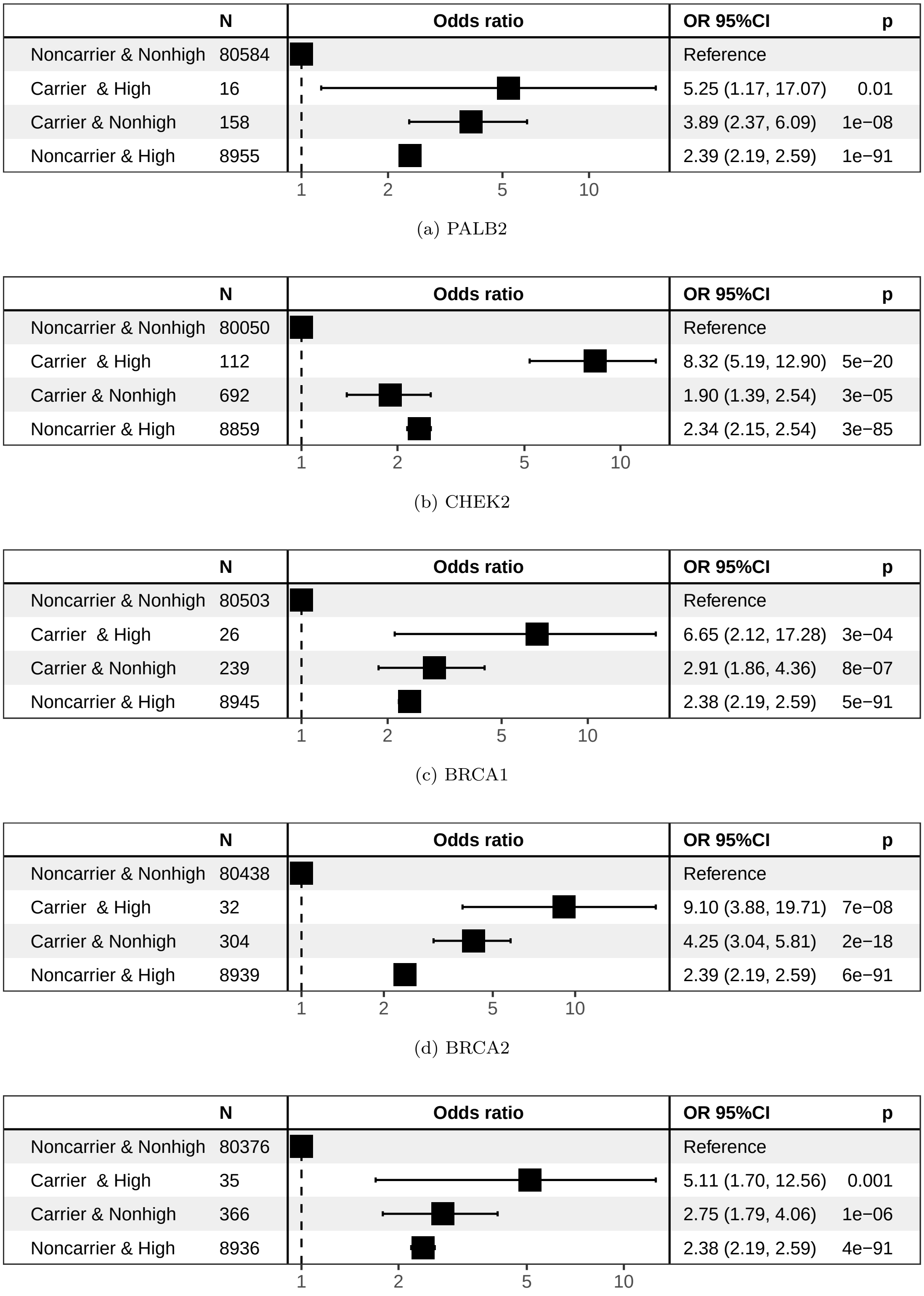

| | N | OR 95%CI | p |
|---|---|---|---|
| Noncarrier & Nonhigh | 80584 | Reference | |
| Carrier & High | 16 | 5.25 (1.17, 17.07) | 0.01 |
| Carrier & Nonhigh | 158 | 3.89 (2.37, 6.09) | 1e−08 |
| Noncarrier & High | 8955 | 2.39 (2.19, 2.59) | 1e−91 |

(a) PALB2

| | N | OR 95%CI | p |
|---|---|---|---|
| Noncarrier & Nonhigh | 80050 | Reference | |
| Carrier & High | 112 | 8.32 (5.19, 12.90) | 5e−20 |
| Carrier & Nonhigh | 692 | 1.90 (1.39, 2.54) | 3e−05 |
| Noncarrier & High | 8859 | 2.34 (2.15, 2.54) | 3e−85 |

(b) CHEK2

| | N | OR 95%CI | p |
|---|---|---|---|
| Noncarrier & Nonhigh | 80503 | Reference | |
| Carrier & High | 26 | 6.65 (2.12, 17.28) | 3e−04 |
| Carrier & Nonhigh | 239 | 2.91 (1.86, 4.36) | 8e−07 |
| Noncarrier & High | 8945 | 2.38 (2.19, 2.59) | 5e−91 |

(c) BRCA1

| | N | OR 95%CI | p |
|---|---|---|---|
| Noncarrier & Nonhigh | 80438 | Reference | |
| Carrier & High | 32 | 9.10 (3.88, 19.71) | 7e−08 |
| Carrier & Nonhigh | 304 | 4.25 (3.04, 5.81) | 2e−18 |
| Noncarrier & High | 8939 | 2.39 (2.19, 2.59) | 6e−91 |

(d) BRCA2

| | N | OR 95%CI | p |
|---|---|---|---|
| Noncarrier & Nonhigh | 80376 | Reference | |
| Carrier & High | 35 | 5.11 (1.70, 12.56) | 0.001 |
| Carrier & Nonhigh | 366 | 2.75 (1.79, 4.06) | 1e−06 |
| Noncarrier & High | 8936 | 2.38 (2.19, 2.59) | 4e−91 |

(e) ATM

Figure S6: Breast cancer odds ratio (OR) among individuals stratified for presence of high-impact variants in each gene and PRS.

| | N | OR 95%CI | p |
|---|---|---|---|
| NonCarrier and Non−HighPRS | 66337 | Reference | |
| Carrier and HighPRS | 68 | 6.26 (2.40, 13.45) | 2e−05 |
| Carrier and Non−HighPRS | 662 | 1.51 (0.86, 2.44) | 0.1 |
| NonCarrier and HighPRS | 7376 | 2.56 (2.23, 2.92) | 2e−42 |

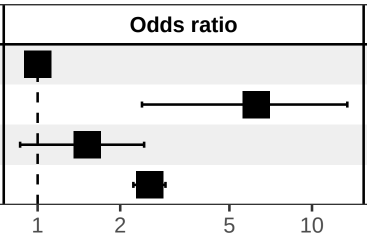


(a) CHEK2

| | N | OR 95%CI | p |
|---|---|---|---|
| NonCarrier and Non−HighPRS | 66746 | Reference | |
| Carrier and HighPRS | 32 | 6.16 (1.46, 17.57) | 0.003 |
| Carrier and Non−HighPRS | 253 | 2.01 (0.90, 3.89) | 0.059 |
| NonCarrier and HighPRS | 7412 | 2.57 (2.25, 2.94) | 2e−43 |

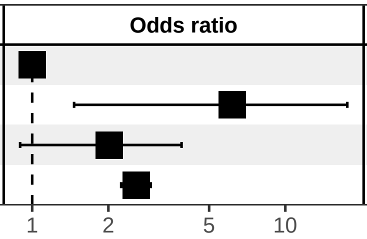


(b) ATM

| | N | OR 95%CI | p |
|---|---|---|---|
| NonCarrier and Non−HighPRS | 66765 | Reference | |
| Carrier and HighPRS | 21 | 6.92 (1.09, 24.38) | 0.01 |
| Carrier and Non−HighPRS | 234 | 0.53 (0.09, 1.66) | 0.37 |
| NonCarrier and HighPRS | 7423 | 2.56 (2.24, 2.93) | 3e−43 |

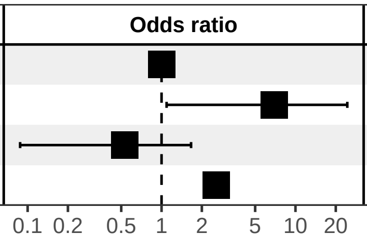


(c) BRCA1

| | N | OR 95%CI | p |
|---|---|---|---|
| NonCarrier and Non−HighPRS | 66741 | Reference | |
| Carrier and HighPRS | 25 | 5.35 (0.85, 18.32) | 0.02 |
| Carrier and Non−HighPRS | 258 | 1.77 (0.75, 3.49) | 0.14 |
| NonCarrier and HighPRS | 7419 | 2.58 (2.25, 2.94) | 1e−43 |

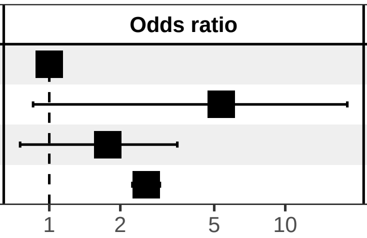


(d) BRCA2

| | N | OR 95%CI | p |
|---|---|---|---|
| NonCarrier and Non−HighPRS | 66828 | Reference | |
| Carrier and HighPRS | 60 | 10.60 (4.61, 21.30) | 9e−10 |
| Carrier and Non−HighPRS | 171 | 4.37 (2.22, 7.73) | 3e−06 |
| NonCarrier and HighPRS | 7384 | 2.54 (2.22, 2.91) | 8e−42 |

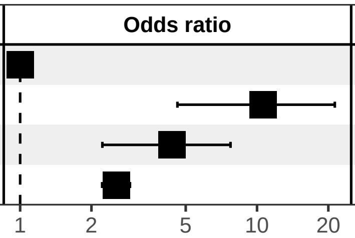


(e) HOXB13

Figure S7: Prostate cancer OR among individuals stratified for presence of high-impact variants in each gene and PRS.

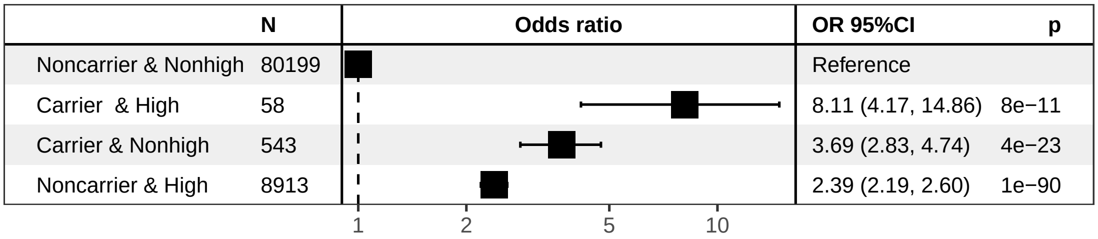


(a) high risk variants (BRCA1, BRCA2)

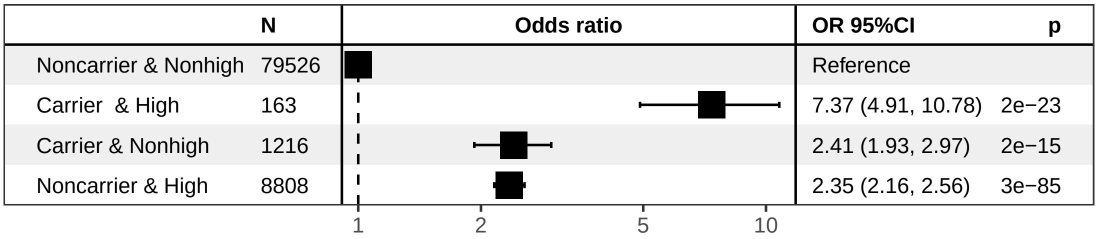


(b) intermediate risk variants (PALB2,CHEK2,ATM)

Figure S8: Breast cancer OR among individuals stratified for presence of high-impact variants carrier in intermediate and high risk variants and PRS.

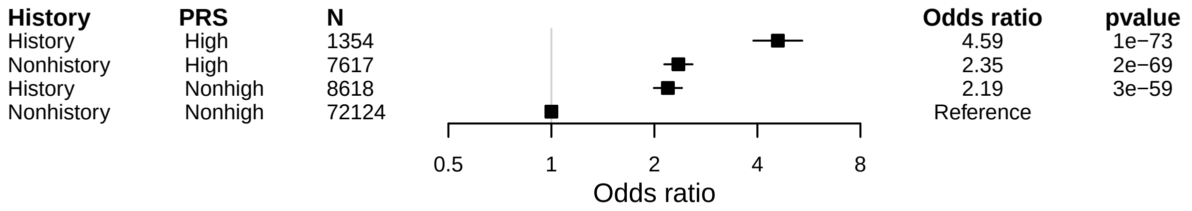


Figure S9: Breast cancer OR among individuals stratified for presence of a first-degree family history with breast cancer and PRS.

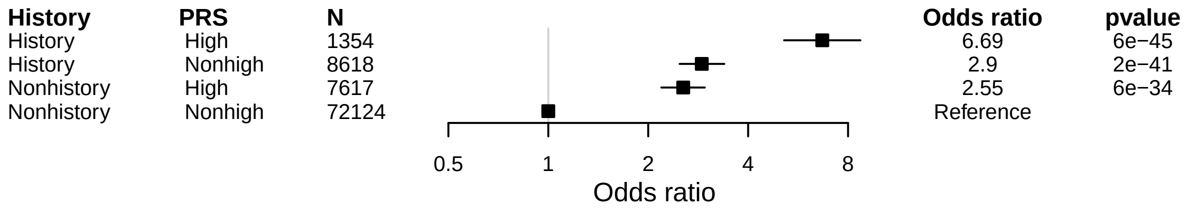


Figure S10: Prostate cancer OR among individuals stratified for presence of a first-degree family history with prostate cancer and PRS.

Table S1: Characteristics of participants by breast cancer status.

| | Cases | Controls |
|---|---|---|
| Participants, N | 3739 | 85974 |
| Age*, mean (SD) | 52.23 (7.57) | 56.83 (8.35) |
| Family history of breast cancer, n (%) | 762 (20.38) | 9210 (10.71) |

* Age: age at onset for cases, and age at recruitment for controls

Table S2: Characteristics of participants by prostate cancer status.

| | Cases | Controls |
|---|---|---|
| Participants, N | 1332 | 73111 |
| Age*, mean (SD) | 60.35 (4.44) | 57.02 (8.68) |
| Family history of prostate cancer, n (%) | 273 (20.5) | 5689 (7.78) |

* Age: age at onset for cases, and age at recruitment for controls
